# Automated molecular testing of saliva for SARS-CoV-2 detection

**DOI:** 10.1101/2020.08.11.20170613

**Authors:** Nancy Matic, Tanya Lawson, Gordon Ritchie, Aleksandra Stefanovic, Victor Leung, Sylvie Champagne, Marc G. Romney, Christopher F. Lowe

## Abstract

With surging global demand for increased SARS-CoV-2 testing capacity, clinical laboratories seek automated, high-throughput molecular solutions, particularly for specimen types which do not rely upon supply of specialized collection devices or viral transport media (VTM). Saliva was evaluated as a diagnostic specimen for SARS-CoV-2 using the cobas® SARS-CoV-2 Test on the cobas® 6800 instrument. Saliva specimens submitted from various patient populations under investigation for COVID-19 from March-July 2020 were processed in the laboratory with sterile phosphate-buffered saline in a 1:2 dilution and vortexed with glass beads. The processed saliva samples were tested using a commercial assay for detection of the SARS-CoV-2 E gene (LightMix®) in comparison to the cobas® SARS-CoV-2 Test. 22/64 (34.4%) of the saliva samples were positive for SARS-CoV-2. Positive and negative concordance between the LightMix® and cobas® assays were 100%. There was no cross-contamination of samples observed on the cobas® 6800. The overall invalid rate for saliva on the cobas® 6800 (1/128, 0.78%) was similar to the baseline invalid rate observed for nasopharyngeal swabs/VTM and plasma samples. Saliva is a feasible specimen type for SARS-CoV-2 testing on the cobas® 6800, with potential to improve turnaround time and enhance testing capacity.

## Introduction

The global public health response to the COVID-19 pandemic highlighted the critical need for diagnostic testing which is sustainable, practical, rapid and scalable(1). With an increasing worldwide demand for SARS-CoV-2 molecular testing, supply-chain issues for high-quality, flocked nasopharyngeal (NP) swabs have created significant challenges for testing capacity in clinical and public health laboratories. Alternate specimen types, such as saliva, have been reported in some studies to have nearly comparable sensitivity to nasopharyngeal swabs for the detection of SARS-CoV-2, and may be an appropriate supplemental or alternate diagnostic specimen(2-4). Although a variety of methods for saliva collection have been described(2, 5, 6), we have previously shown the utility of testing saliva in the absence of transport media, which enables a simple collection technique that avoids the introduction of potential inhibitors(7, 8) and dependence on supply of specialized saliva collection devices.

In addition to potentially obviating supply shortages, saliva has been increasingly described as a useful sample for the diagnosis of COVID-19 to overcome certain pre-analytical collection challenges. Flocked NP swabs have been the preferred specimen type due to established sensitivity, but may occasionally result in false-negative test results due to poor specimen collection quality(9) or timing of testing relative to symptom onset(10-12). Lower respiratory tract specimens such as a bronchoalveolar lavage are often obtained from severely ill patients, but require aerosol-generating medical procedures and thus have the potential for aerosolization and transmission of SARS-CoV-2. Furthermore, only a minority of patients with COVID-19 are able to produce expectorated sputum(13). Saliva is a convenient alternate sample to collect for SARS-CoV-2 detection, particularly for patients with high clinical suspicion for COVID-19 but repeatedly negative test results by NP swabs(14, 15) or for individuals unwilling or unable to tolerate NP swab collection.

Within our clinical laboratory, processing and testing of saliva is currently a manual process requiring extraction (MagNA Pure Compact or MagNA Pure 96, Roche Molecular Diagnostics, Pleasanton, CA) followed by amplification (LightCycler 480, Roche). Post-analytical reporting into the electronic medical records system is also a manual process. As a result, capacity for saliva testing in our laboratory is limited, with delays in turnaround time compared to nasopharyngeal swabs which are processed entirely on the automated cobas® 6800 (Roche). We sought to evaluate the potential utilization of the cobas® 6800 for SARS-CoV-2 detection from saliva.

## Materials and Methods

From March-July 2020, saliva was ordered by clinicians from both hospitalized and ambulatory patients for the diagnosis of COVID-19. Briefly, ≥1mL of saliva was collected in a sterile screw-top container (Starplex Scientific Inc., Etobicoke, Canada) without the addition of transport media, and then processed in the virology laboratory with phosphate-buffered saline (PBS) in a 1:2 dilution and glass beads. Samples were tested using the LightMix® ModularDx SARS-CoV (COVID19) E-gene assay (TIB Molbiol, Berlin, Germany), with use of the MagNA Pure Compact or MagNA Pure 96 and LightCycler 480. The remaining volume of processed saliva samples was stored at −70°C. Processed saliva samples were then tested with the cobas® SARS-CoV-2 Test (Roche Molecular Diagnostics, Laval, QC) on the cobas® 6800. Prior to cobas® SARS-CoV-2 testing, a software upgrade (Assay Specific Analysis Package [ASAP]) was required on the cobas® 6800 to prevent viscous specimens from mistakenly being interpreted as clotted/invalid by the instrument. Samples known to be positive for SARS-CoV-2 based on results from the LightMix® assay were alternated with negative samples in a checkerboard pattern on the 96-well processing plate, and tested in duplicate.

## Results

A total of 64 clinical saliva samples were included and tested in duplicate for SARS-CoV-2 on the cobas® 6800. Twenty-two (34.4%) of the samples were known to be positive for SARS-CoV-2 based on prior results from the LightMix® assay, and 42 (65.6%) had no detectable SARS-CoV-2. Compared to the LightMix® assay, positive percent agreement and negative percent agreement on the cobas® 6800 were 100%. The median cycle threshold (Ct) values for detection of the Envelope (E) gene were comparable between the cobas® 6800 (28.82, interquartile range [IQR] 7.29) and the LightMix® assay (26.63, IQR 7.61) (**Figure 1**). No carryover or cross-contamination of samples was observed, even with strongly positive samples (Ct values 13 to 24) directly adjacent to negative samples. One saliva sample produced an error which was reported by the cobas 6800® instrument as ‘Invalid’; this sample was successfully tested on the duplicate run. In total, the observed error rate for saliva samples tested on the cobas® 6800 was 0.78% (1/128).

**Figure 1:**
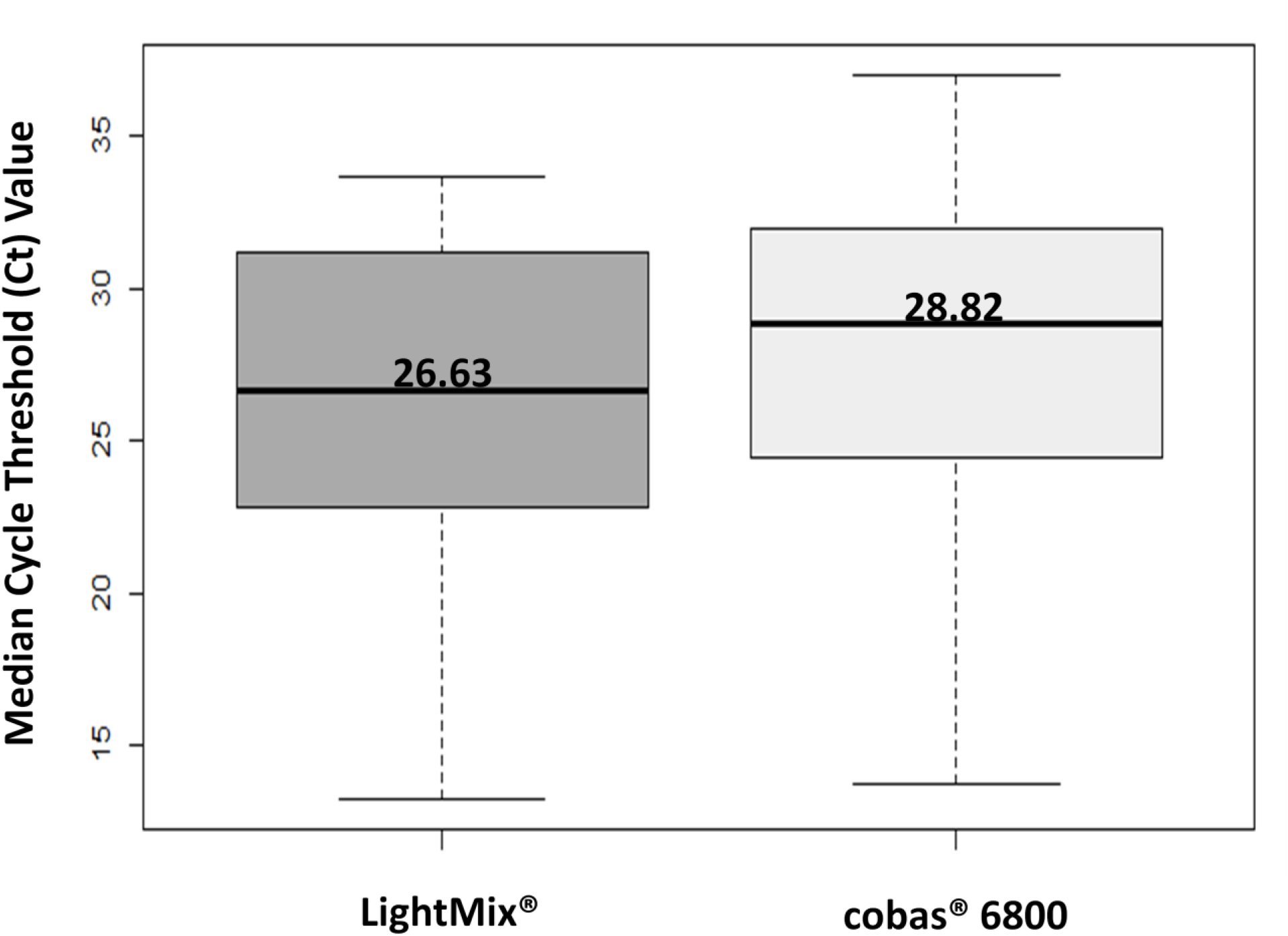
No significant difference in median cycle threshold (Ct) values for the Envelope (E) gene was observed in saliva samples positive for SARS-CoV-2, using the LightMix® ModularDx SARS-CoV (COVID19) E-gene assay (TIB Molbiol, Berlin, Germany) and the cobas® SARS-CoV-2 Test (Roche Molecular Diagnostics, Laval, QC) on the cobas® 6800 platform.

## Discussion

In our evaluation of the cobas® 6800 for saliva testing, we demonstrated complete concordance in comparison to the LightMix® assay, which is the current assay utilized in our laboratory for clinical testing of specimens other than NP swabs such as saliva.

Previous studies have evaluated the feasibility of saliva for SARS-CoV-2 detection on commercial instruments such as the Cepheid GeneXpert® System(16) and cobas® 8800(12). However, there are technical challenges associated with processing saliva for automated, high-throughput testing. As a highly viscous sample compared to viral transport media, there is potential for pipetting errors or instrument contamination. In our laboratory, saliva is pre-processed with sterile PBS and glass beads in order to decrease the viscosity of the sample. The samples utilized in the study were from a variety of clinical situations, including long-term care residents, inpatients at a tertiary care hospital, and outpatient contacts of known COVID-19 patients; these samples would be broadly representative of saliva ordered and collected for clinical testing in the future. Reassuringly, testing with this specimen type did not result in a significant number of invalid results (1/128, 0.78%), and was comparable to the rate of invalid results observed in our laboratory from nasopharyngeal swabs with the SARS-CoV-2 Test (0.20%) and plasma used with other cobas® 6800 assays (0.30%) (unpublished data). We also assessed the potential for carryover during pipetting on the cobas® by testing positive samples alternated with negative samples in a checkerboard pattern, and did not identify any cross-contamination of samples.

This study is limited by the retrospective testing of stored saliva samples. Clinical testing was performed in real time on the LightMix® assay, and samples were stored for subsequent testing on the cobas® after the software had been upgraded. Although testing was not performed in parallel, there was high concordance of testing despite sample storage. Furthermore, testing of retrospective samples was necessary to ensure a sufficient number of saliva samples with detectable SARS-CoV-2 RNA were included.

Due to limited remnant sample volume in some cases, five of the twenty-two saliva samples positive for SARS-CoV-2 (22.7%) underwent additional dilution with sterile PBS in order to test on the cobas® 6800. The Ct values in these cases cannot be directly compared to those from the LightMix® assay as a measure of assay performance; however, the dilution factor in these cases (1:3 to 1:5) would have had a negligible effect on the Ct values, and the aim of this study was to assess the technical feasibility of saliva samples for automated, high-throughput SARS-CoV-2 testing on the cobas® 6800.

Transitioning to an automated platform for saliva testing is critical for enhancing SARS-CoV-2 testing capacity, particularly in preparation for a potential resurgence of COVID-19 cases or mass testing of defined populations. Automated testing reduces errors in the pre-analytical and post-analytical phases, and improves turnaround time by enabling saliva to be processed on multiple runs per day (including overnight testing). Our evaluation confirmed the feasibility of saliva as a suitable specimen type for SARS-CoV-2 testing on the cobas® 6800 platform.

## Data Availability

This article is a focused, brief report with the majority of data already presented in the manuscript.

## Acknowledgements

We are grateful to our medical laboratory technologists who are highly committed to patient care and laboratory quality improvement.

This study was supported by Roche Diagnostics for a software upgrade of the cobas® 6800, but no financial compensation was provided. Roche Diagnostics was not involved in the study design, implementation nor analysis of the data.

This study received approval from the Providence Health Care Research Ethics Board.

